# Patterns of Reported Infection and Reinfection of SARS-CoV-2 in England

**DOI:** 10.1101/2022.06.13.22276316

**Authors:** Matt J. Keeling

## Abstract

One of the key features of any infectious disease is whether infection generates long-lasting immunity or whether repeated reinfection is common. In the former, the long-term dynamics are driven by the birth of susceptible individuals while in the latter the dynamics are governed by the speed of waning immunity. Between these two extremes a range of scenarios is possible. During the early waves of SARS-CoV-2, the underlying paradigm was for long-lasting immunity, but more recent data and in particular the 2022 Omicron waves have shown that reinfection can be relatively common. Here we investigate reported SARS-CoV-2 cases in England, partitioning the data into four main waves, and consider the temporal distribution of first and second reports of infection. We show that a simple low-dimensional statistical model of random (but scaled) reinfection captures much of the observed dynamics, with the value of this scaling, *k*, providing information of underlying epidemiological patterns. We conclude that there is considerable heterogeneity in risk of reporting reinfection by wave, age-group and location. The high levels of reinfection in the Omicron wave (we estimate that 18% of all Omicron cases had been previously infected, although not necessarily previously reported infection) point to reinfection events dominating future COVID-19 dynamics.

## 1 Introduction

The pattern of SARS-CoV-2 cases in England and elsewhere can be conceptualised as a series of waves, often associated with the emergence of a new variant. In England, there have been four main waves: the first from January to July 2020 was due to the Wildtype variant, the second from August 2020 to May 2021 was due to the Wildtype variant followed by the Alpha variant, the third from May to December 2021 was attributable to the Delta variant, while the fourth wave has been driven by the Omicron variant (with two sub-waves due to sub-types BA.1 from December 2021 to March 2022, and BA.2 from March 2022 onwards). The scale of these waves is dependent on the characteristics of each variant [1] (with each variant having a higher transmission potential than the last and by immune escape for the Omicron variant), population mixing [2] (impacted by control measures and precautionary behaviour driven by perceived risk) and population immunity [3], with the latter being the most relevant for the long-term dynamics.

From the start of the epidemic in January 2020 until 14th April 2022 (our chosen end date due to changes in national testing policy in England [4]), there were 18.3 million reported infections, equivalent to 33% of the English population. This is likely to be a considerable underestimate of the true number of infections, especially in the early waves when testing was far more limited. During the first wave (from the first reported cases in England in late January 2020 until 15th July 2020) there were around 250,000 reported cases - whereas the first REACT study estimated around 3.4 million people had been infected by this time [5]. In the second wave, testing became more wide-spread with national testing centres and postal tests available. From April 2021 to April 2022, covering much of the third and fourth waves, free Lateral Flow Device (LFD) tests were freely available in England, and many groups were encouraged to perform regular tests, hence during these later times we may expect the ratio of reported cases to infection to increase. Despite the under-reporting, the study of identified cases and in particular individuals who test positive on two (or more) occasions is extremely valuable.

During the first three waves (associated with Wildtype, Alpha and Delta variants) reported reinfections were rare; less that 1% of cases before December 2021 were in individuals that had previously reported infection. Reinfections increased dramatically during the Omicron waves, with around 10% of cases from January to April 2022 having previously reported infection.

## 2 Methods

England has had multiple methods of testing for SARS-CoV-2 infection operating in parallel. These can be split into: Pillar 1 testing - generally PCR-based in hospitals and care homes; and Pillar 2 testing - generally at home or in testing centres [6]. The nature of Pillar 2 testing has changed over the course of the pandemic, with such testing largely absent during the first wave. Subsequently, during the second and third waves of the pandemic PCR testing was advised for all symptomatic infections and as a follow-up to positive LFD tests, with PCR comprising 95% of all positive Pillar 2 tests in 2020 and 2021. From early 2022 onward LFD tests dominate. Here, we do not distinguish between the type of test, but use episode number and an anonymised unique identifier to link first and second infections - we combine reported cases from both Pillar 1 and Pillar 2 test, LFD and PCR positive tests without discriminating. In particular, we label *C*_*t,T*_ to be the number of individual reported cases that first test positive on day *t* and then subsequently on day *T* (*T* > *t* + 90); we also define *P*_*t*_ as the total number of all positive tests on day *t*, irrespective of whether they are a first or subsequent infection. We note that there is a 90-day threshold imposed on the data we receive, such that a new episode is only defined if it is more the 90 days since the last positive test. This 90-day threshold ensures that long-duration infections with multiple positive tests are not counted as repeat infections [7]; however this threshold could hide some rapid reinfection or might not exclude all long-duration infections - we will see that secondary episodes at 90 days are relative common.

As such *C*_*i,j*_ is a matrix of values, where *i* and *j* convey information on the likely variants while the value of *C* informs on the time-varying testing behaviour and the level of cross-protection. Examining all the positive tests in England from late January 2020 to 14 April 2022, we find that approximately 95% are first reports, 5% are second reports and only 0.064% (approximately 12,000 cases) are third or subsequent reports of infection. For this reason we restrict our attention to first and second reports only, which simplifies the interaction between variants and reporting interval that needs to be considered.

We partition the period January 2020 to April 2022 into four distinct waves and note the dominant variant(s): Wave 1, Wildtype (from the first case on 30th January - 15th July 2020); Wave 2, Wildtype and Alpha (16th July 2020 - 30th April 2021); Wave 3, Delta (1st May - 12th December 2021); and Wave 4, Omicron, both BA.1 and BA.2 (from 12th December 2021 onwards). We compare the data on first and second reported cases (*C*_*t,T*_) at times *t* and *T*, to a simple model (*M*_*t,T*_) in which reinfection is a random process, that could have wave-specific scaling:

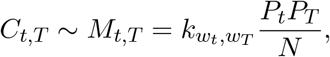

where *N* is the population size (approximately 56 million for England), *P*_*t*_ is the number of cases at time *t* (we note that *P*_*t*_ ≫ Σ_*T*_ *C*_*t,T*_ as *P* refers to all cases, whereas *C* are only individuals that report two or more infections) and *w*_*t*_ refers to the wave number at a particular time point. Under this formulation, *k* would be 1 for a homogeneous infection obeying the SIS paradigm with no immunity and equal risk of reporting all cases, so that a proportion *P*_*t*_*/N* of the *P*_*T*_ reported cases are expected to have reported previous infection at time *t*. Immunity will act to reduce *k*; we expect *k* ∝ (1 − *ρ*) where *ρ* informs about degree of cross-protection afforded by previous infection (or alternatively 1 − *ρ* is the the relative risk of reinfection); however, heterogeneity in the risk of infection and reporting within the population will act to increase *k* - as the effective population size experiencing and reporting infection will be smaller.

We consider two particular forms of *k*, estimated such that *M*_*t,T*_ is a good fit to *C*_*t,T*_, in terms of minimising the root-mean square difference. Firstly, a homogeneous model where just two values of *k* are used, an initial value for the first three waves when reinfection was rare, and a second higher value during the fourth Omicron wave when the degree of protection was lower and reinfection was more common:

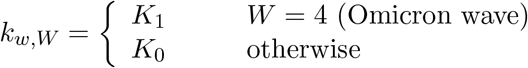

Alternatively, we can compute a different *k*_*w,W*_ for every pair of waves (noting that *w* ≤ *W*).

To delve into the interactions within and between waves in more detail, we consider the different pairs of waves and the delay, *d*, between first and second infections in waves *w* and *W* as captured by

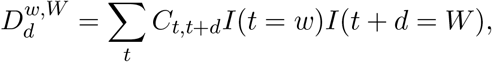

where *I* is used to identify if a time period is associated with a particular wave. The simple model, *M*_*t,T*_ (making either the homogeneous and variant-specific assumptions for *k*) can also be used to generate a similar function:

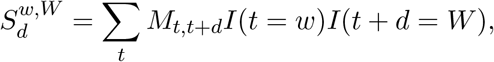

capturing the expected pattern of reported reinfection events over time.

We therefore use the estimated values of *k* to understand the general patterns of reinfection between waves, noting that both greater risk of re-infection or greater heterogeneity can lead to an increased value of *k*, such that we cannot uniquely attribute every change in *k* to a direct epidemiological cause - although the relative level of reported cases provides some guidance. We use the comparison between *D* and *S* to explore the finer structure within each wave, and to assess longer-term trends.

## 3 Results

Figures 1 and S1 graphically compare the data (*C*_*t,T*_) to the simple models of reinfection (*M*_*t,T*_). Focusing on the homogeneous assumption for *k* (Fig. 1) we estimate that *K*_0_ = 0.184 (CI 0.183−0.185) and *K*_1_ = 0.648 (CI 0.646 − 0.649) which taken together explain 51% and 85% of the variance in reinfection data in the pre-Omicron (waves 1-3) and Omicron periods (wave 4) respectively. (That is, var(*C*_*t,T*_ − *M*_*t,T*_)*/*var(*C*_*t,T*_) is 0.51 for *T* in the first three waves and 0.85 for *T* in the fourth wave; with much of the remaining variance due to the relatively small and integer nature of *C*, whereas *M* is represents a continuous expectation.) The top panel of Figure 1 is simply the number of reported cases over time (*P*_*t*_), with reinfections in dark grey; the bottom panel is the number of first cases of individuals who report a secondary infection (Σ_*T*_ *C*_*t,T*_, plotted against *t*), while the right-hand panel is the number of secondary reported infections (Σ_*s*_ *C*_*s,t*,_ plotted against *t*). We note that due to the much larger number of reinfection events for Omicron (presumably caused by a lower *ρ* value and less cross-protection from previous infection, reflected in *K*_1_ > *K*_0_), we have been forced to use two different scales for the pre-Omicron and Omicron periods, as captured by the top and bottom axes.

**Fig. 1:**
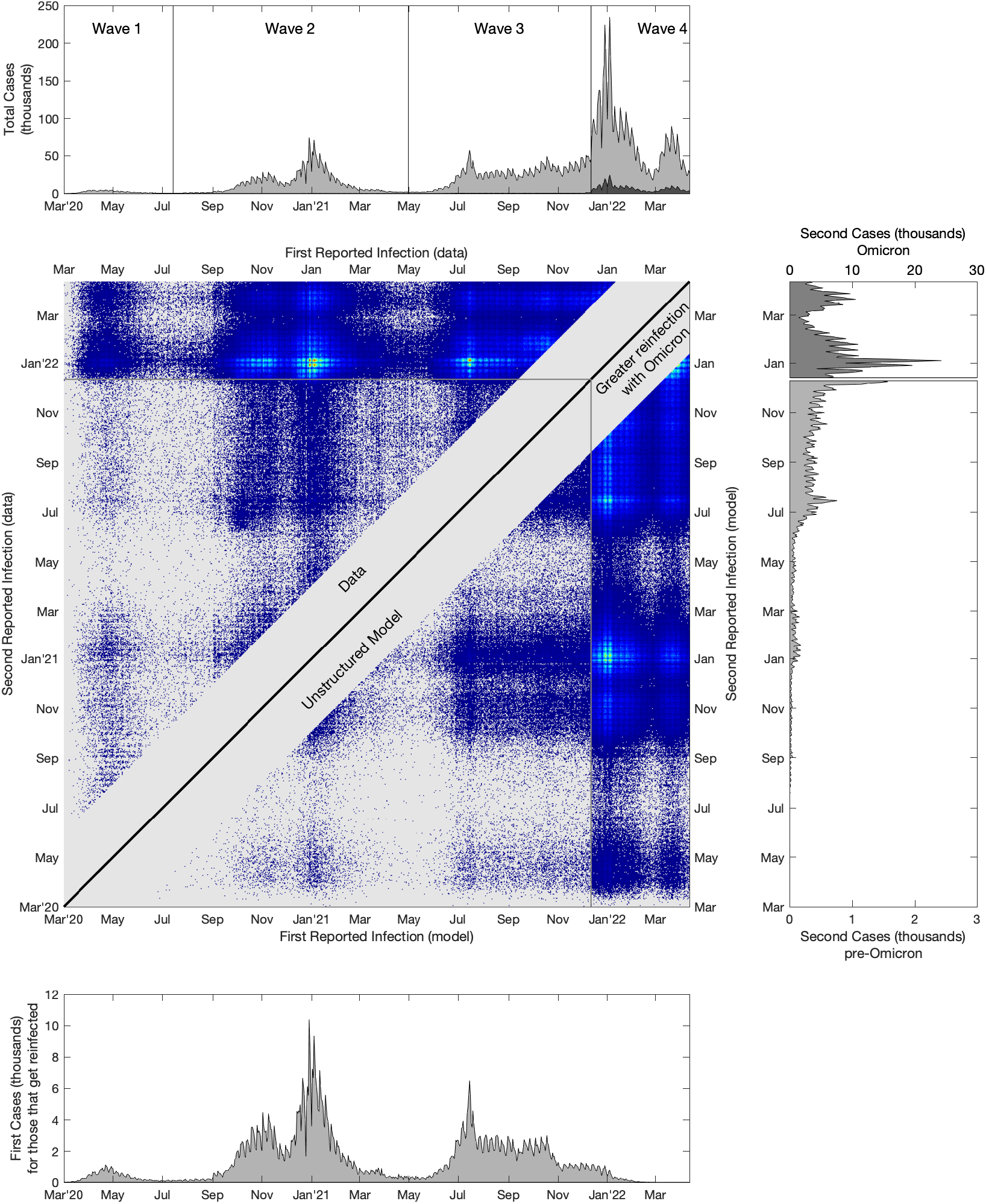
A comparison of reported first and second infections in England to the simple homogeneous assumption model. The central panel shows reinfection data (*C*_*t,T*_ upper-left triangle) and the corresponding model fit (in particular we show a Poisson sample of *M*_*t,T*_ in the lower-right triangle, to allow a better visual comparison when *M*_*t,T*_ is low), brighter colours correspond to more reported reinfections, while grey is zero reinfections in the data or model. The upper panel shows the total number of reported cases (pale grey) and the number of second reported cases (dark grey); the lower panel shows the number of reported first cases for those that report twice, while the right-hand panel shows the second reports - note that second reported cases during the Omicron wave (after 12th December 2021) are plotted on a different scale for clarity.

Finally, the middle panel shows the data on reinfections (*C*_*t,T*_ upper-left triangle) and the simple model (*M*_*t,T*_ lower-right triangular) - we note that the bottom and right panels are therefore the appropriate projections of the sum of the matrix of data (*C*_*t,T*_). From inspection, it is clear that reinfection is far more common in the Omicron waves (as exemplified by *K*_1_ being at least three times larger than *K*_0_), and that the simple model captures the bulk patterns of reinfection, even with this homogeneous set of assumptions for *k*.

Figure S1 shows similar results for the model in which *k* is wave specific, with the four main UK waves (associated with Wildtype, Wildtype/Alpha, Delta and Omicron). This greater heterogeneity in *k* explains more of the variance in *C*_*t,T*_ : 56% and 90% for the pre-Omicron and Omicron periods respectively. The relatively small improvement in the pre-Omicron results (compared to a single *k* value) is because *C*_*t,T*_ is dominated by the single combination of reinfections in wave 3 (Delta variant) after an initial infection in wave 2.

We now consider the data transformed to examine the separation between first and second cases for each wave (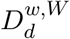, Fig. 2), which in plotted in the same upper triangular form as the data in Figure 1. To this data we add lines as produced from the simple model 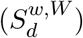 with both the homogeneous (dashed line) and wave-specific (solid line) assumptions for *k*.

**Fig. 2:**
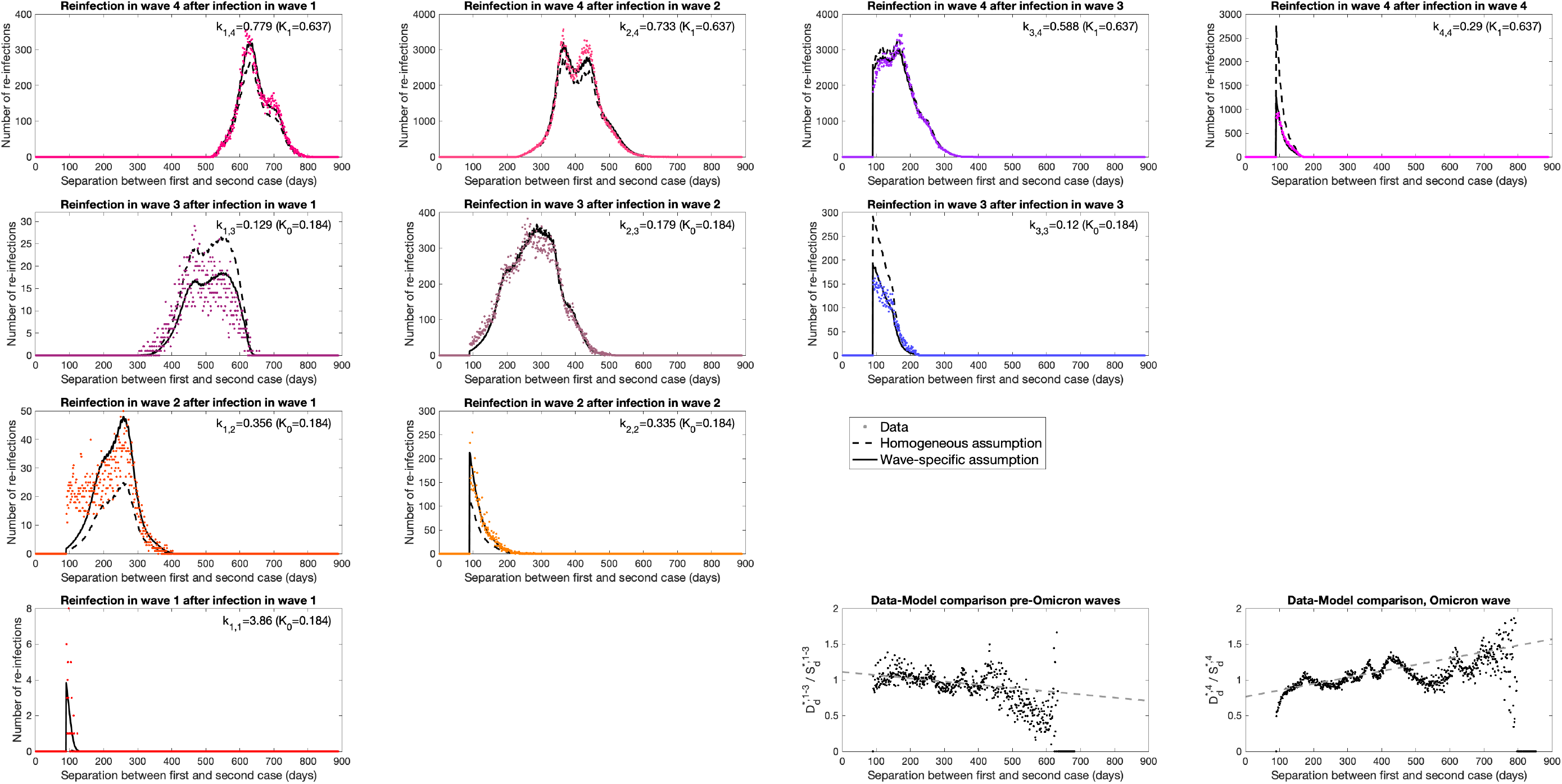
Wave-wave interactions for reported reinfections. Each panel in the upper-left triangle shows the number of reported cases with a given delay between the first and second report (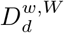, coloured dots), separated by the associated waves. For each, we also show the same numbers based on the simple model with both the wave-specific (solid lines) and homogeneous (dashed lines) assumptions for *k*. Note, to enable visual inspection of the empirical data compared to the simple model, we have used different y-axis ranges across subplots. The two panels in the bottom right perform a comparison of data 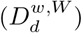 and model (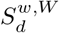, using the homogeneous assumption) when the second cases are from the pre-Omicron (*W* ≤ 3) and Omicron (*W* = 4) waves respectively, enabling us to detect temporal trends, shown as a linear fit to the points.

For Wildtype-Wildtype reinfections in wave 1 (Fig. 2, bottom left), the value of *k* for the variant-specific model is large (approximately 3.86), suggesting that reinfection with wildtype is much more common than expected from case numbers; we attribute this to the extreme heterogeneity in reporting leading to a much smaller effective population size, as only a small fraction of the population was likely to be tested (in general only those severely ill and requiring hospital treatment). (All values of *k* for the variant-specific assumption together with 95% confidence intervals, are given in Table 1.) Reinfections in wave 2 show a similar pattern to our observations for wave 1 (Fig. 2, row 3), where the wave-specific model *k* is larger (0.335 and 0.356) than in the homogeneous model (0.184), but far smaller than for wave 1 due to increased population-level testing. The model generally captures the timing of wave 2 reinfections but substantially underestimates rapid reinfection (days 90-150) in the transition between wave 1 and wave 2 (which would correspond to Wildtype-Wildtype reinfections), although the total number of such reinfections is relatively small.

**Table 1:** The scaling parameter 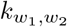 that generates the best fit between the simple model and the data; 95% confidence intervals are also given assuming that the data represent a Poisson sample with the model as the mean. We maintain the same upper-triangular structure as seen in Fig. 1 and Fig. 2.

| Wave<br>second case | Wave, first case |  |  |  |
| --- | --- | --- | --- | --- |
|  | 1 | 2 | 3 | 4 |
| 4 | 0.766 (0.757 - 0.775) | 0.735 (0.733 - 0.737) | 0.591 (0.589 - 0.593) | 0.235 (0.231 - 0.240) |
| 3 | 0.129 (0.124 - 0.132) | 0.179 (0.177 - 0.180) | 0.120 (0.117 - 0.122) | - |
| 2 | 0.356 (0.347 - 0.365) | 0.335 (0.327 - 0.342) | - | - |
| 1 | 3.86 (3.50 - 4.22) | - | - | - |

Delta variant reinfections during wave 3 (Fig. 2, row 2) show far more agreement between the simple model and the data. Secondary cases with Delta after a primary case in wave 2 (Fig. 2, row 2 column 2) numerically dominate, and for such reports the wave-specific and homogeneous models generate similar *k* values (*k*_2,3_ = 0.179 compared to *K*_0_ = 0.184). The fact that *k* is less than one is a reflection of the protection afforded by the primary case; however, it does not suggest that the degree of cross-protection, *ρ*, is around 82% (1 − *k*_2,3_), as due to heterogeneity in infection and reporting risk, the effective population size is likely to be far smaller than the true population size. We note a slightly reduced level of reinfection (for Delta reinfection after wave 1 cases, although the numbers are again small, and for Delta after Delta) in the data and variant-specific model compared to the homogeneous model, which may be a signal of greater cross-protection.

Lastly, for reinfections in the Omicron (fourth) wave, (Fig. 2, top row) the data and models are in relatively good agreement, with both models generating similar fits. The estimated values of *k* are all relatively large (0.591 to 0.766) which we attribute to high rates of reinfections due to limited cross protection. The exception to this pattern is Omicron-Omicron pairs where the variant-specific *k* is 0.235, suggesting that there is a higher degree of protection against the same variant. Considering the temporal pattern in more detail, we note that *k* decreases from initial cases during wave 1 through to initial cases with Delta in wave 3, hinting a decline in cross-protection over time. In addition, within each subplot there is a tendency for the model to slightly overestimate the number of most recent reinfections or underestimate the number of longer duration reinfections - which again suggests a slight decline in cross-protection over time.

To explore this temporal aspect in more detail, we compare the data, 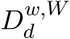, with the model, 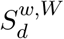 (by plotting *D*_*d*_*/S*_*d*_ against *d*). We contrast when the second infection, *W*, occurs during any of the first three waves, with when reinfection occurs in the fourth Omicron wave (Fig. 2, bottom right). We perform this comparison using the homogeneous model (*k* = *K*_0_ pre-Omicron or *k* = *K*_1_ for Omicron) as this removes the temporal component that is inherent within the wave-specific assumptions. In the pre-Omicron period we observe a slight but statistically significant (*p* < 10^−7^) decline in *D*_*d*_*/S*_*d*_ with the separation between cases, *d*; whereas during Omicron the trend is stronger and in the opposite direction (*p* < 10^−100^). Therefore during the Omicron wave, the model (with homogeneous assumption for *k*) tends to underestimate the number of recent reported reinfections, and consequently overestimate the number of longer separation reinfections; the trend of more infections at longer separations may be a signal of waning immunity.

We can use the analysis of these reported reinfection patterns to generate estimates of the total level of reinfection. For the four waves considered here, we estimate a reporting rate of 7.3% (6.6-8.0%) in wave 1, 32% (28-38%) in wave 2, 56% (46-65%) in wave 3 and 31% (25-40%) in wave 4, using the fitted Warwick model [8–10]. This agrees with primary measurements: estimates from the REACT-2 study [11] lead to a 7.5% (7.2-7.9%) reporting rate in wave 1 (based on measurements that 6.0% (5.8-6.1%) were infected in the first wave [5]), while cumulative incidence estimates from the ONS study [12] lead to a 38% (36-41%) and 42% (39-46%) reporting rate for waves 2 and 3 respectively (Fig. S3). Using this level of reporting through time, and assuming that reported cases are representative of all infections, we can estimate the true level of reinfection (after 90-days) over the course of the pandemic. For wave 1, the amount of reinfection is low at around 0.6% (0.5-0.7%), for waves 2 and 3 this increases to 2.5% (2.2-2.8%) and 2.3% (2.0-2.5%) respectively, but for wave 4 and the Omicron variant this increases to 18% (16-20%). This is strongly suggestive that the future dynamics of SARS-CoV-2 will be contingent on reinfections within the population, although the extent to which these are mild or severe infections will be driven by the characteristics of new variants that emerge.

Finally, we perform the same analysis but on subsets of the population; in particular we partition the population by age and location (Fig. 3). We calculate *k* in the pre-omicron waves (Fig. 3, blue) and Omicron wave (Fig. 3, red). Both show distinct deviation from the population average values (*K*_0_ = 0.184 and *K*_1_ = 0.648), with patterns that differ between pre-Omicron and Omicron waves. Pre-Omicron (January 2020 to December 2021, Fig. 3 left) the scaling parameter *k* is large for those over 60, those between 20 and 30, and those under 5 (there are more reported reinfections than expected from population averages); for the Omicron wave, it is only the 7-11 year olds and 20-22 year olds that are appreciably greater than *K*_1_, with the 70-80 year olds showing a substantially reduced level of reported reinfection compared to population-level expectations. Spatially, in the pre-Omicron waves, there is greater reporting reinfection than expected in Cornwall, Devon and East Anglia; whereas during the Omicron wave, we observe that it is predominantly Southern Central England that has lower than expected reinfection.

**Fig. 3:**
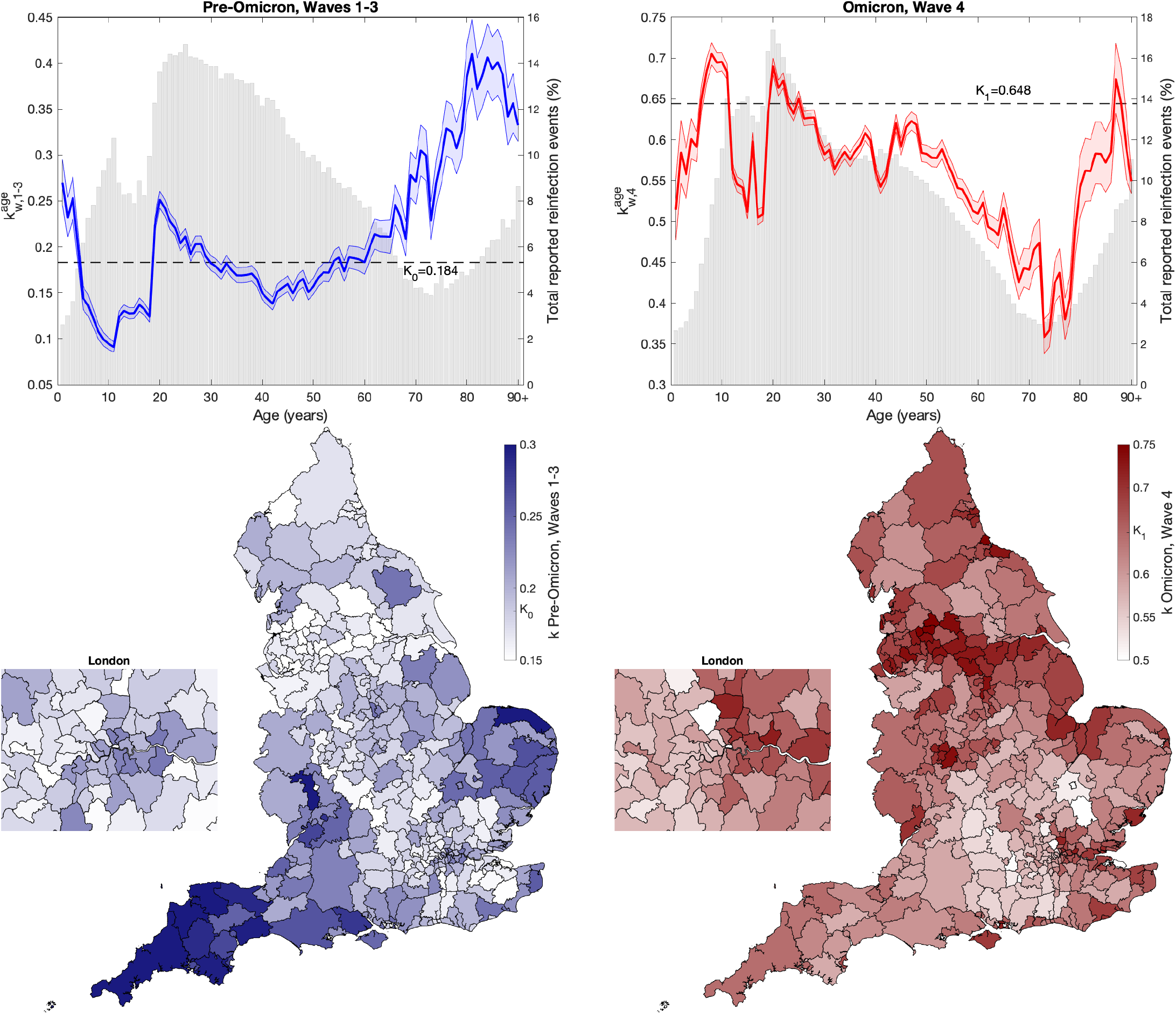
The values of the scaling *k* when partitioning the data by age or spatial location, in comparison to the homogeneous assumption *k* = *K*_0_ or *K*_1_. Top row shows the mean value of *k* in one-year age groups (from 0 to 90+), together with 95% confidence intervals, the left-hand figure (blue) is for waves 1-3 pre-Omicron, while the right-hand figure (red) is for Omicron wave 4. The grey bars (and right-hand y-axis) show the percentage of all reported cases that are reinfections (Σ_*t,T*_ *C*_*t,T*_ */*Σ_*T*_ *P*_*T*_). The lower panels are when the data is partitioned by Lower Tier Local Authority (LTLA) of which there are 317 in England; darker colours refer to higher *k* values, with *K*_0_ (pre-Omicron waves national average value for *k*) and *K*_1_ (Omicron wave national average value for *k*) shown on the colorbar.

## 4 Discussion

We have passed the point in the pandemic where the overwhelming majority of people in England have already been infected, with estimates of 70.7% (90% credible interval: 66.0-75.6%) infected by 11th February 2022 [12] - although many of these will have been mild or asymptomatic infections and not reported. The ability of SARS-CoV-2 to reinfect individuals is a key factor in both its persistence and control in the longer-term and the transition to endemicity [13, 14]. Here, using detailed individual-level case reports for England from the start of the pandemic (January 2020) to 14th April 2022, we consider the pattern of reported cases and the number of reported reinfection events. By far the most striking patterns are the massive increase in reported reinfection during the Omicron waves, and the general agreement between the data and a simple model of random reinfection.

The fact that the random model fits the data so well, even using the homogeneous assumption for *k* (which involves the fitting of just two scaling quantities), strongly suggests that there are at most sparse immunological signals in this data. The data are well captured by a model in which reinfections occur randomly in a subset of the population (accounting for protection from past infection, heterogeneous risk of infection and heterogeneous likelihood of testing in a single parameter, *k*), with only a minimal impact of the temporal separation between infections. For second reported infections with Omicron during wave 4, there is a slight signal of waning immunity - reporting reinfection when the initial infection was over 18 months ago is 35% more likely than reporting reinfection when the initial infection was less than 6 months ago given the relative abundance of reported cases over these periods (Fig. 2, bottom-right). However, before Omicron this trend was reversed (although weaker) with shorter separations being relatively more likely (Fig. 2, bottom column 3).

The scaling parameter *k* is seen to vary substantially between waves, and between ages and spatial locations. *k* is a combination of two factors - the degree of protection offered by past infection (which reduces *k*) and the degree of population heterogeneity (which increases *k*); in general it is impossible to disentangle these two elements, from *k* alone; however combining changes in *k* with changes in reported cases and reinfections offers some insights. The large jump in the number of reported reinfections in the Omicron wave suggests that the associated increase in *k* is primarily associated with a reduction in cross protection. In contrast, we believe that the high value in the first wave (*k*_1,1_ = 3.86) reflects the narrow subset of the population that would be tested for SARS-CoV-2 infection, thereby reducing the effective population size. In particular, given the same pattern of infection, the value of *k* will double if only half the population would report an infection, as the effective population size is halved. This basic concept can be extended, with heterogeneity in risk of infection or the chance of reporting leading to a increase in *k*. Moreover, vaccination is likely to increase *k* if both first and second case are post vaccination roll-out, as this changes the heterogeneity in risk across the population (those unvaccinated have a higher chance of infection).

The age and spatial data require more care to interpret. In the Omicron wave (Fig. 3, top-right), the shape of *k* closely follows the pattern of reported reinfections, suggesting that the low in 70-80 years old is due to increased cross protection (or behavioural changes) following previous infection; whereas the high *k* value and low reinfection probability in 7-11 year olds hints at highly heterogeneous levels of infection and reporting - potentially due to a subset of this age-group regularly self-testing. Similarly, the patterns pre-Omicron (Fig. 3, top-left), where the elderly population have high *k* and relatively low reported reinfections, are suggestive of dynamics driven by population heterogeneity, especially in the elderly.

There are some important caveats to this work. Firstly, we are only dealing with reported cases; this either requires symptomatic illness followed by testing (and reporting) or periodic testing to detect mild or asymptomatic infection. This is important for four main reasons. The pattern of testing has changed throughout the pandemic as different forms of testing have become available, in the first wave testing was of severely ill patients only and using PCR, whereas from April 2021 to April 2022 free lateral flow device (LFD) tests were available nationally. There is also a strong age-bias in testing with younger individuals more likely to test regularly and therefore detect asymptomatic infections. We note that there is evidence that for individuals that are infected more than once, the most common pattern is for just one severe episode with other infections being mild or asymptomatic [15, 16] - this means that reporting of multiple infections is likely to be an underestimate unless there is regular asymptomatic testing. Only positive tests separated by 90 days are recorded as a new reported episode, meaning that shorter separations between reinfection events are not included and long-duration persistent infections, although rare, may be recorded as multiple episodes.

Secondly, we have subdivided the timeline into four periods, roughly corresponding to the four main waves; while waves 1, 3 and 4 are dominated by Wildtype, Delta and Omicron variants respectively, wave 2 is a mixture of Wildtype and Alpha. An alternative characterisation, splitting wave 2 into Wildtype and Alpha dominated periods (Fig. S2), does not greatly improve the fit; in particular there are problems capturing reported reinfections with Wildtype variants in wave 2 after initial infection in wave 1. Our definition of waves is not rigorous, but we generally transition at the point of minimum reported infections, such that the precise timing of the transition does not affect the bulk proprieties reported; the only exception to this rule is the transition between waves 3 and 4, which is aligned with the dominance of the Omicron variant. In addition, we do not split the two Omicron waves attributable to BA.1 (during December 2021, January and February 2022) and BA.2 (mainly during March and April 2022), this is due to the short time-scales involved - the 90 day minimum separation between reported infections means that we are left with relatively little discriminatory power and certainly no prospect of have recorded two infections within the BA.2 time period.

Finally, the pattern of reported reinfection events as a function of the time between them (Fig. 2) is largely driven by the timing of the infection waves. Thus while it is intuitively tempting to seek a immunological explanation of the observed pattern of declining reinfection events with separation for cases within the same wave (diagonal subplots in Fig. 2), the decline in reinfection events is a facet of the waves of infection and is echoed in the behaviour of the simple model. When infection has largely peaked in a single wave, the most common separation between any two infection events is short with long separations corresponding to rarer infection events at the start and end of the wave.

These results indicate that reported reinfection events are very much driven by the contemporary and historic pattern of reported cases, with factors such as waning immunity playing a limited role. Heterogeneity is most profound in the difference between pre-Omicron (before December 2021) and Omicron (after December 2021) waves, and the differences between age-groups. Taken together, our results suggest that periodic reinfection is highly likely especially in an environment where new variants are constantly emerging. We predict that the Omicron wave (15th December 2021 to April 2022) was associated with 18% (16-20%) of cases being reinfections. This pattern of reinfections aligns most closely with an SIRS-paradigm, such that we may expect seasonal waves of infection driven by waning immunity and virus evolution.

## Data Availability

Data on cases were obtained from the COVID-19 Hospitalisation in England Surveillance System (CHESS) data set that collects detailed data on patients infected with COVID-19. These data contain confidential information, with public data deposition non-permissible for socioeconomic reasons. The CHESS data resides with the National Health Service (www.nhs.gov.uk). The ethics of the use of these data for these purposes was agreed by Public Health England with the Governments SPI-M(O) / SAGE committees. More aggregate data is freely available from the UK Coronavirus dashboard: https://coronavirus.data.gov.uk/

## 5 Supplementary Figures

**Fig. S1:**
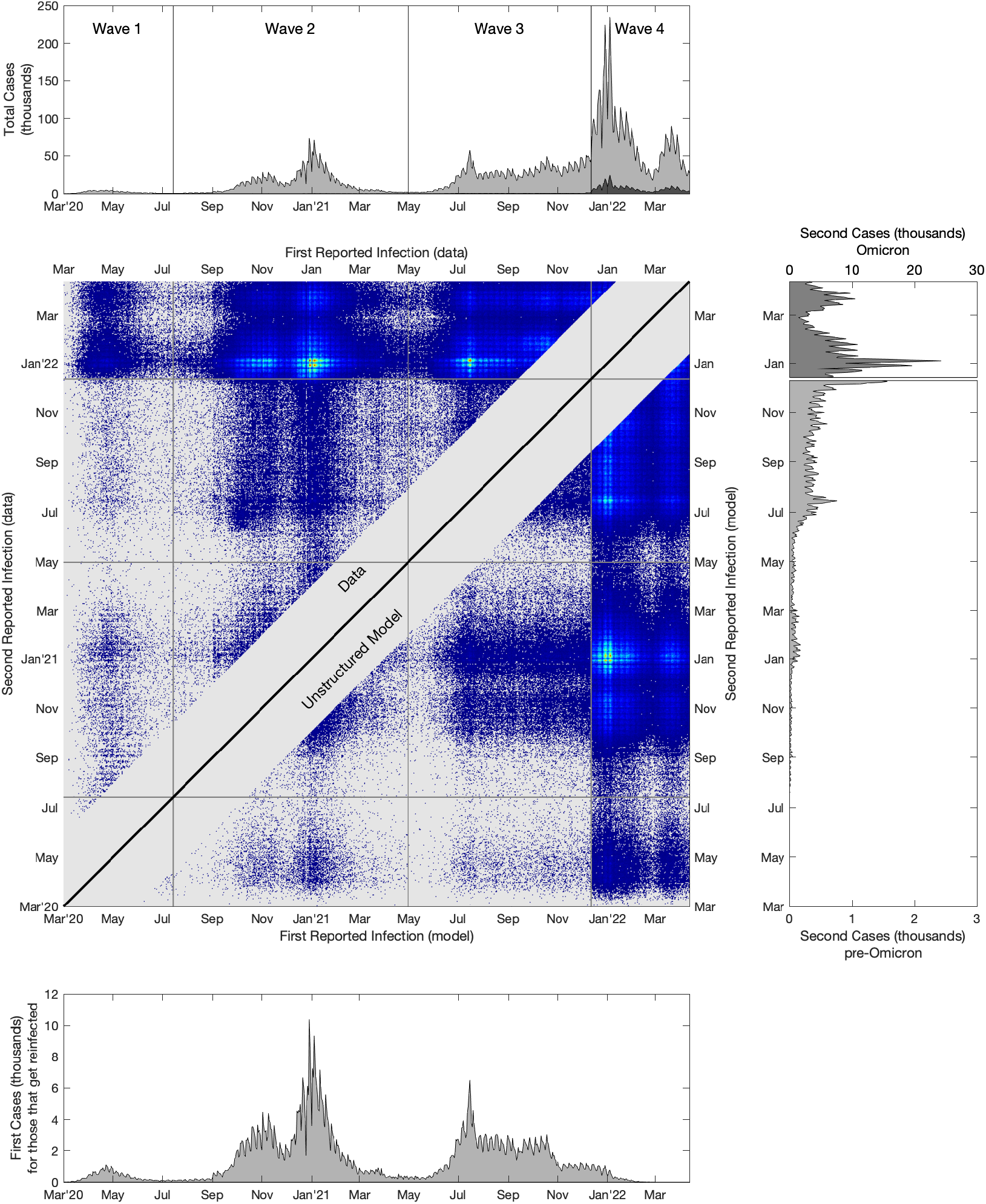
A comparison of reported first and second infections in England to the model where *k* is wave specific. The central panel shows reinfection data (*C*_*t,T*_ upper-left triangle) and the corresponding model fit (in particular we show a Poisson sample of *M*_*t,T*_ in the lower-right triangle, to allow a better visual comparison when *M*_*t,T*_ is low), brighter colours correspond to more reported reinfections, while grey is zero reinfections in the data or model. The upper panel shows the total number of reported cases (pale grey) and the number of second reported cases (dark grey); the lower panel shows the number of reported first cases for those that report twice, while the right-hand panel shows the second reports - note that second reported cases during the Omicron wave (after 12th December 2021) are plotted on a different scale for clarity.

**Fig. S2:**
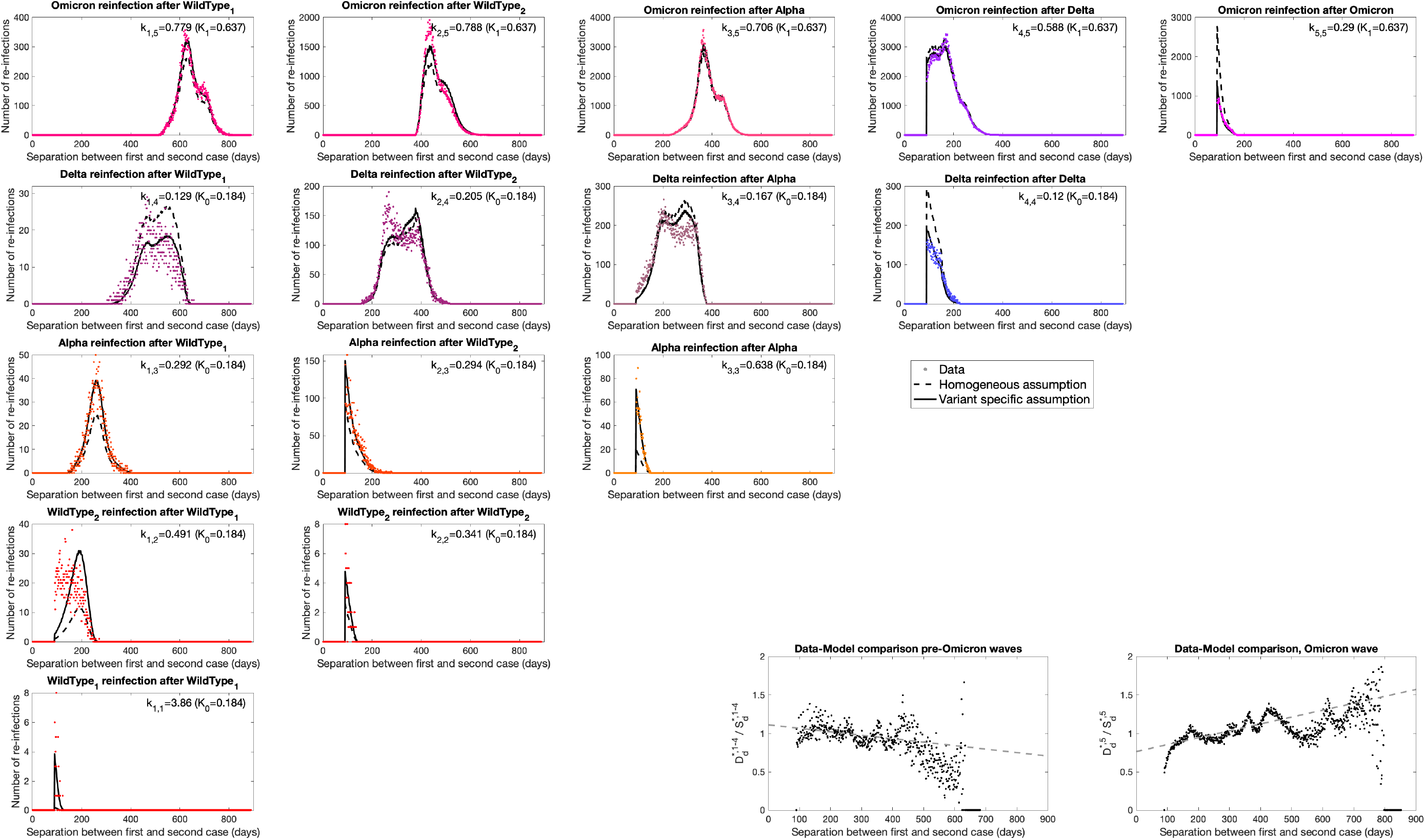
Variant-variant interactions for reported reinfections, by splitting wave 2 (July 2020 to May 2021) into a Wildtype and Alpha variant component. Each panel in the upper-left triangle shows the number of reported cases with a given delay between the first and second report (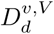, coloured dots), separated by the associated variants. For each, we also show the same numbers based on the simple model with both a variant-specific (solid lines) and homogeneous (dashed lines) assumptions for *k*. Unfortunately, determining the exact variant would require genotyping of all samples; while the ability to detect the S-gene is ready used as a marker of the switch between variants (with Wildtype, Delta and Omicron BA.2 being S-gene positive, and Alpha and Omicron BA.1 being S-gene negative) this only applies to individuals that are PCR tested through the TachPath system. The two panels in the bottom right perform a comparison of data and model (using the homogeneous assumption) when the second cases are from the pre-Omicron and Omicron waves respectively, enabling us to detect temporal trends - which are comparable to those in Fig. 2.

**Fig. S3:**
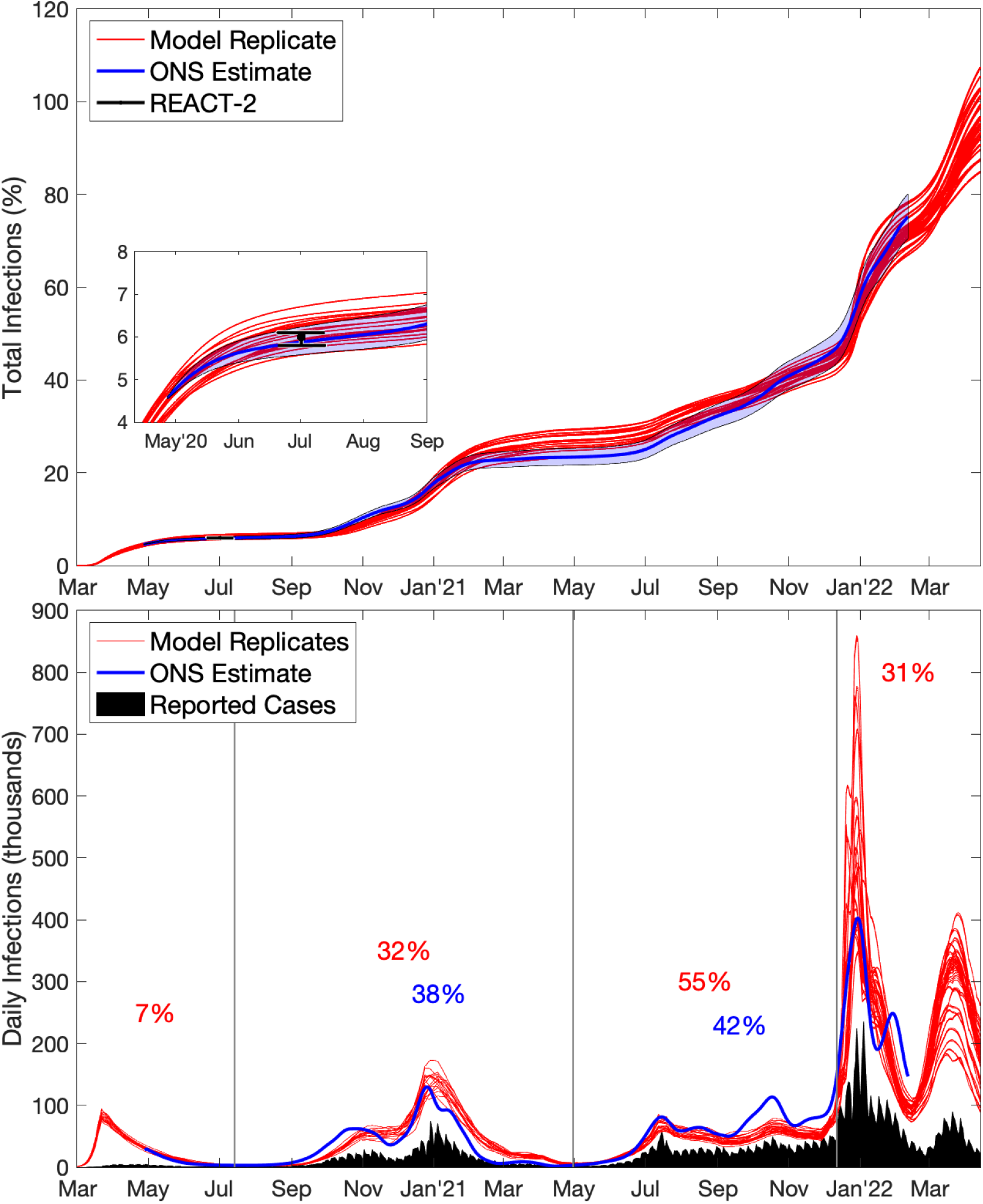
Comparison between different methods of capturing the number of infections. Top: The total cumulative percentage of the population infected over time, showing three different results: (1) total number of infections from the Warwick model [8–10] which has been continually fitted to epidemiological data since the beginning of the epidemic and accounts for regional differences, age-structure and vaccination (red showing, 100 trajectories); the proportion of the population that has been infected from the ONS survey [12] (with the first point on 27th April 2020 increased to match model replicates, blue with 95% credible intervals); and the number of antibody positive individuals in June/July 2020 from the REACT-2 survey [5] (black point and error bars). Bottom: Estimated number of daily infections, from: reported daily cases including reinfection events (black) which forms a lower bound; model replicates including reinfection events (red) and an ONS estimate computed as the rate of change in the total number ever infected from the top graph (blue). The ONS estimate should be a reliable estimate of daily infections for the first three waves when reinfection is uncommon. For each wave (where possible) we display the fraction of infections (from either the Warwick model or the ONS estimate) that are reported as cases.

## Acknowledgements

I would like to thank Ed Hill, Louise Dyson and Mike Tildesley for their extremely helpful comments on the early versions of this work.

## Ethical Considerations

Data from the CHESS and SARI databases were supplied after anonymisation under strict data protection protocols agreed between the University of Warwick and Public Health England. The ethics of the use of these data for these purposes was agreed by Public Health England with the Government’s SPI-M(O) / SAGE committees.

## Funding

MJK was supported through the JUNIPER modelling consortium [grant number MR/V038613/1] and the National Institute for Health Research (NIHR) [Policy Research Programme, Mathematical and Economic Modelling for Vaccination and Immunisation Evaluation, and Emergency Response; NIHR200411]. MJK is affiliated to the National Institute for Health Research Health Protection Research Unit (NIHR HPRU) in Gastrointestinal Infections at University of Liverpool in partnership with UK Health Security Agency (UKHSA), in collaboration with University of Warwick. MJK is also affiliated to the National Institute for Health Research Health Protection Research Unit (NIHR HPRU) in Genomics and Enabling Data at University of Warwick in partnership with UK Health Security Agency (UKHSA). The views expressed are those of the author(s) and not necessarily those of the NHS, the NIHR, the Department of Health and Social Care or UK Health Security Agency.

## Competing interests

All authors declare that they have no competing interests.

## Notes

### Competing Interest Statement

The authors have declared no competing interest.

### Author Declarations

Data from the CHESS and SARI databases were supplied after anonymisation under strict data protection protocols agreed between the University of Warwick and Public Health England. The ethics of the use of these data for these purposes was agreed by Public Health England with the Governments SPI-M(O) / SAGE committees.

